# Perceptions of public about Blood Transfusion in Allied hospital Faisalabad: A qualitative study

**DOI:** 10.1101/2025.06.24.25330083

**Authors:** Noman Aslam, Haleema Shafiq, Aiza Noor Shahid, Mahnoor, Ayesha Ayub

**Affiliations:** Final year MBBS in Faisalabad Medical University; (Supervisor)/Head of Health Professions Education and Research Department in Faisalabad Medical University

**Keywords:** Blood transfusion, Blood donation, Perceptions, Public

## Abstract

**Background:** Blood donation and transfusion practices are very crucial for a health care system. This study aims to check the perceptions of Public about blood donation and transfusion practices in Allied Hospital, Faisalabad (Pakistan).

**Study Design and Methods:** The study used a qualitative exploratory design, conducting 20 semi-structured interviews with patients and attendees of the surgical and gynaecology unit at Allied Hospital, Faisalabad, from April to June 2023. Using Random sampling technique, data was collected from both donors and non-donors. Then, data was analysed using inductive thematic analysis, which involved identifying patterns and themes within the data.

**Results:** Initial analysis revealed various codes which were ultimately reduced to five main themes with their subthemes namely 1. Personal Factors with sub-themes (1a) Blood donation priorities, (1b) Family influence and Physical Attributes of donor, (1c) Socio economic status, (1d) Gender roles, 2. Consequences of transfusion and Donation; sub-themes (2a) Allergic reaction, (2b) Weight changes after Blood Donation, 3. Motivating Factors for Blood Donation; sub-themes (3a) Health Benefits, (3b) Humanitarian Factors, 4. Restriction Factors for Blood Donation; sub-themes (4a) Selling of Donated Blood, (4b) Substandard Blood Bank, (4c) Wasting of Donated Blood and 5. Awareness regarding blood donation.

**Conclusion:** This study shows that religion, personal beliefs, family and friends, and socioeconomic status influence willingness of a person to donate. However, low understanding of benefits and risks is prevalent among Pakistanis, highlighting the need for counselling and health education activities.

## INTRODUCTION

Blood Donation and transfusion practices are a very important part of holistic medicine. Each year, hundreds and thousands of patients need blood and blood-based products but maintaining an adequate blood supply is still a challenge due to its limited shelf-life.Presence of adequate blood donors is recommended worldwide to combat this problem [1].World Health Organization (WHO) and International Federation of Red Cross and Red Crescent Societies both classify three types of blood donors: paid donors, replacement donors, and volunteer donors. The WHO also encourages all nations to develop their own systems, structures, and procedures for efficiently handling of donated blood.Long-term, this will contribute to keeping blood readily available whenever it is required [2].Globally, an effective healthcare delivery system is recognized by safe blood units that should be transfused under various clinical scenarios. These may include acute hemorrhage due to road traffic accidents (RTA), in a post-partum and antepartum hemorrhage, or anemic patients. Proper administration of blood and blood-based products could be life-saving under such conditions [3].The average blood donation rate in developed countries is 31.5 donations per 1000 people. In comparison to the above value, 16.4 donations per 1000 people are seen in upper middle-income countries, 6.6 donations per 1000 people in lower-middle-income countries, and 5.0 donations per 1000 people in low-income countries [4].In era of technology, developing countries including Pakistan facing challenge of low blood supplies despite having huge source pools in form of youngsters. According to the Global Database on Blood Safety, 25% of blood donations in Pakistan are from the Voluntary Donors. In Pakistan, the Replacement Donors account for 70% of all donations, while the Paid Donors account for 10% of the country’s blood donating population [5]. Factors responsible for such low blood collection include mismanagement at blood banks, flaunts in supply chains, misperceptions of public, lack of education and awareness about the need of safe blood, and high prevalence of Transfusion Transmitted infections (TTIs) [6]. The supply and safety of blood are major issues in Pakistan. One of the main concerns still remains to be the high prevalence of TTI, which includes HBV, HCV, HIV, syphilis, and malaria [7]. Also, The National Institute of Health (NIH) guidelines for Pakistani blood services generally replicate rules from other nations (primarily developed countries) without taking into account local issues and people’s ground realities [8].Although much work is done throughout the world to assess the donors’ and non-donors’ attitudes towards blood donation, barely anything is done in Pakistan and much effort is required in this field. Otherwise, lack of blood donor motivation and retention measures could obsolete the concept of voluntary donors in near future [9]. Shortly, mismanagement of blood pools will force us to pay debts in the future. The purpose of this study is to access the perception of the Pakistanis about blood and transfusion practices to rule out major factors so that the statistics of blood donation could become better in Pakistan.

## STUDY DESIGN AND METHODS

A Qualitative study design was employed to explore the perceptions of the public regarding blood donation and transfusion practices in Pakistan. The study consisted of 20 semi-structured interviews conducted from April to June 2023 at Allied Hospital, Faisalabad; a Public sector tertiary care hospital in Pakistan. Before conducting the research, ethical approval was taken from the Ethical Review Committee, before initiation of data collection under the No.F.48-ERC/FMU/2022-23/257. As research contains human population so informed consent was taken from every participant before commencement of each interview. It was a written consent which contained their signs and thumb impressions.

Simple Random Sampling tool was used to specify the participants. All the patients and their attendees present in the Gynecology and Surgical unit of Allied Hospital were eligible for participation in study. No discrimination of gender was made at any step. 12 out of 20 participants were males and 8 were females. Informed consent is taken from participants, interviews were recorded on devices as well as noted manually. Semi structured, open-ended questions were used in interviews to allow participants to express a variety of viewpoints and to help our team gather comprehensive data. The interactions during the interviews allowed dynamic exchange of ideas and free-flow discussions about the issues. To maintain the confidentiality and to avoid biasing as far as possible, data transcripts were anonymized. Patients privacy was maintained as no personal information like name and images are used in this article. The method of analysis used for this study was an inductive thematic framework analysis and allowed us to identify multiple themes and sub-themes.

## RESULTS

This study included individuals with diverse demographical characteristics in terms of gender, transfusion status and age as shown in Fig 1 and Fig 2. Participants were only selected from the Surgical and Gynecology unit of Allied Hospital on random basis.After analyzing the data, six themes were generated. Impact of religion on blood donation 2. Motivating Factors for Blood Donation: 2a) Humanitarian Factors, 2b) Health Benefits, 3. Restriction Factors for Blood Donation: 3a) Selling of Donated Blood; 3b) Substandard Blood Banks; 3c) Wasting of Donated Blood; 4. Personal Factors: 4a) Blood donation priorities; 4b) family influence and physical attributes; 4c) socioeconomic status; 4d) low incidence of women in blood donation. 5. Complications of Blood Transfusion: 5a) allergies; 5b) weight changes; 6. Awareness regarding blood donation.

**Figure 1.**
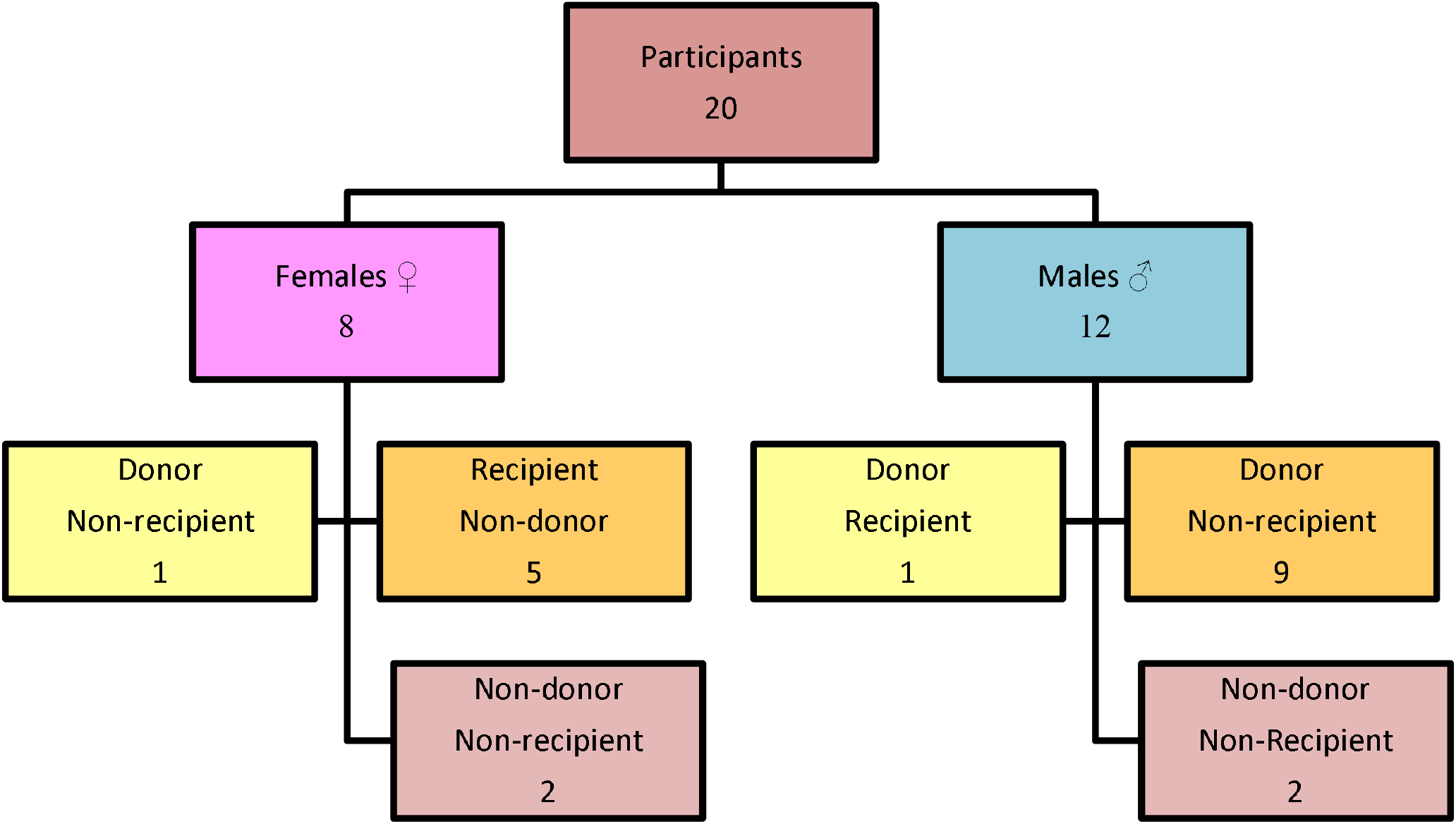
Demographic details of Participants.

**Figure 2.**
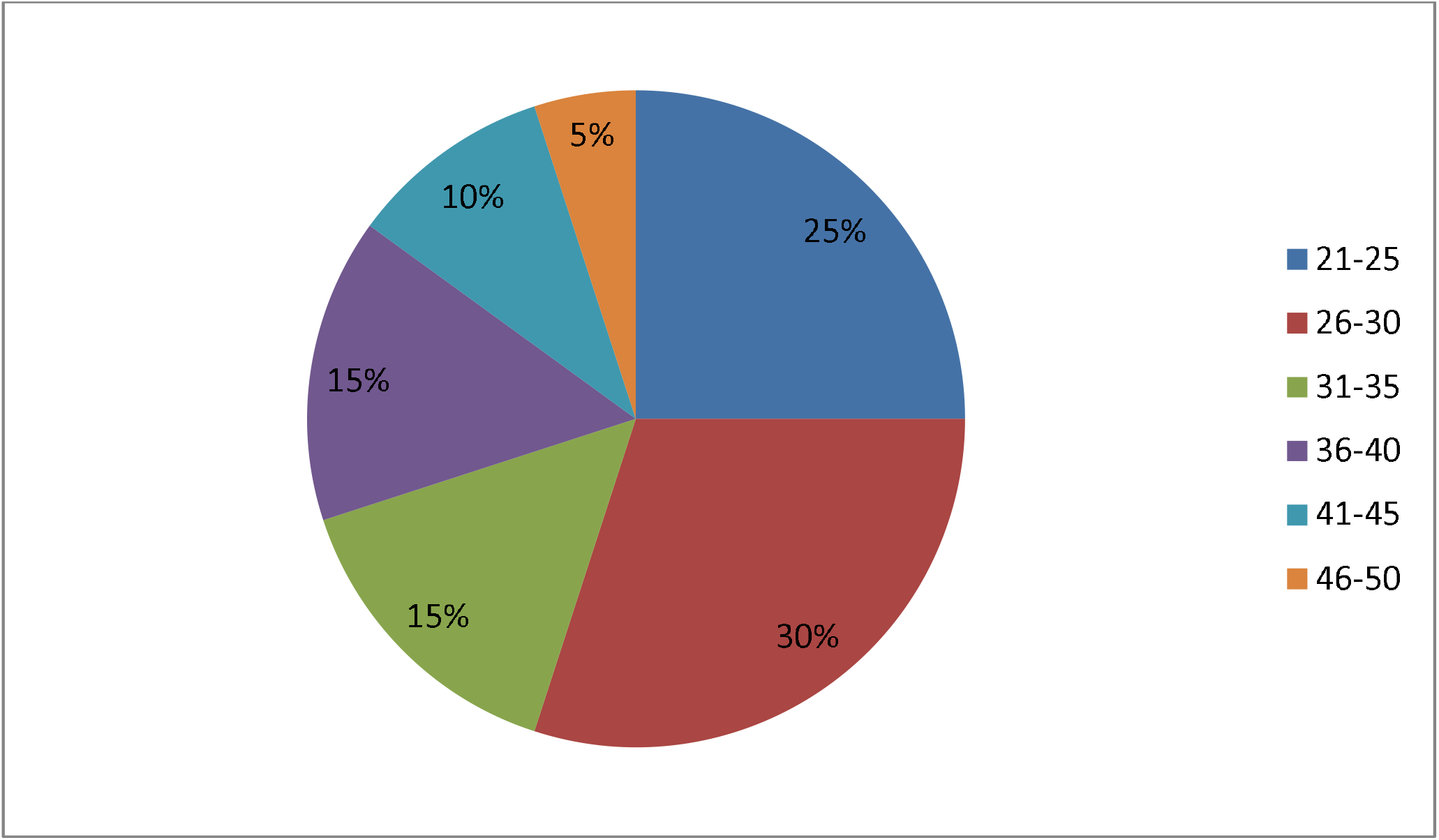
Age distribution of Participants.

1. **Impact of religion on Blood Donation** In this study, all of the twenty participants were Muslims with positive attitudes towards blood donation. Among them, seven narrated donation as “a continuous act of compassion”. Many cited the Hadith: saving one life is equivalent to saving the whole of humanity. To gain clarity, two participants even talked to Islamic scholars and consulted Islamic sources.
2. **Motivating Factors for Blood Donation**
  2a) Humanitarian Factors The majority of participants reasoned altruism as their primary driving force for blood donation. They considered donating blood as a wonderful way to help the individual saying, “Blood Donation has nothing to do with education. It is a matter of humanity.”
  2b) Health Benefits All participants supported blood donation because of its positive effects on health. Five participants said, ‘the body regenerates blood after donation and that the production of fresh blood makes them healthy’. Four interviewees said that they had witnessed their friends and family members donate blood, and after the donation, they were healthy. One of them narrates, “My brother had some kind of allergy in his legs, and after blood donation, his allergy had disappeared and he felt healthy.”
3. **Restriction Factors for Blood Donation**
  3a) Selling of Donated Blood Seven participants had seen donated blood being sold after collection through donation camps while eight had witnessed the same in nearby hospitals. Four only heard about blood selling but never witnessed it. Further, some added that trust in blood camps can only be restored if proper assurance is provided.
  3b) Substandard Blood Banks Participants told that basic facilities like syringes, blood bags, etc. Were not available in blood banks. Some even stated that “There is a lot of corruption in the blood banks, and they give us stored blood, which typically causes allergies in its recipients” .Others suggested that blood banks should screen all diseases properly, as practiced in foreign countries.
  3c) Wasting of Donated Blood In risky operations, doctors demand to arrange blood for emergencies, but such preparation is wasted when not used. Few individuals blamed doctors for the blood waste based on their experience. “Doctors occasionally advise us to arrange blood, and then don’t give blood to our patient after that,” they alleged.
4. **Personal Factors**
  4a) Blood Donation priorities According to this study, people prefer to donate blood in their close circle, including family, friends, relatives, and colleagues. Likewise, people also preferred to receive blood from close relationships. Nine participants stated the reason for accepting blood donations only from family members: “Blood from a stranger can react”. The remaining participants said they could accept donations from strangers only in case of emergencies. Further, the pattern of regular donations was found to be weak in society, as no one in our study was a regular donor or had ever encountered one.
  4b) Family influence & Physical Attributes: The family had influenced a lot in determining the willingness of people to donate blood. Most had supportive families. However, some interviewees claimed that even being capable of donating blood; they were opposed by their mother and wives due to their health concerns. Four respondents were demoralized by family due to fear of becoming weak after donation. They said a donor should be stout and of heavy physique.
  4c) Socio economic status Almost all of our participants came from middle-class backgrounds. Five of them worked on daily wages and could not afford financial compensation after blood donation. Four said, “See the inflation; how will we eat and drink to replenish our blood later on?” One of them even considered the possibility of family emergency for blood, so he found himself unable to make a blood donation.
  4d) Low Incidence of Women in blood donation: In our study, most of the women thought that they were too weak for blood donation. Only one out of eight women was the donor. Three women supported their inability to donate blood as the reason; we are usually blood deficient and comparatively live a difficult life. While four women said,” Women have to face monthly menstrual bleeding and various gynecological surgeries during their lives, so they are physically unable to make blood donation”. One participant said, “Nobody has ever asked me for a donation; if they had, I would have given it. On asking, ten male participants put forward their opinion that women can only donate blood if their hemoglobin level is normal. Two male participants said that they don’t find it ethically appropriate for themselves that their ladies donate blood when they can.
5. **Complications of Blood Transfusion**
  5a) Allergies Five participants reported that blood from strangers had caused allergic reactions. Most used ‘palpitations’, ‘shaking’, ‘elevated blood pressure’, ‘fever’, and ‘skin rashes’ to describe post-transfusion response. The majority quoted unreliable lab tests and blood from smokers as the main causes for the reaction. However two interviewees had witnessed a transfusion reaction even when the donor was a non-smoker. Seven said, “An addict can transfer his or her addiction and all the toxic chemicals into the body of the recipient through blood.”
  5b) Weight changes: Majority who connected blood donation with weight changes stated, “We have heard from our family and seen in friends that blood donation had increased their body weights.”
6. **Awareness regarding blood donation** Almost all participants observed a lack of awareness in Pakistan regarding blood transfusion practices. On asking the best practical way to spread awareness, six chose to include it in school syllabus, four preferred seminars at college and high school level to address youth and rest four suggested that teams should visit home to home to guide about transfusion. The summary of results is given in Table 1.

**Table 1.**
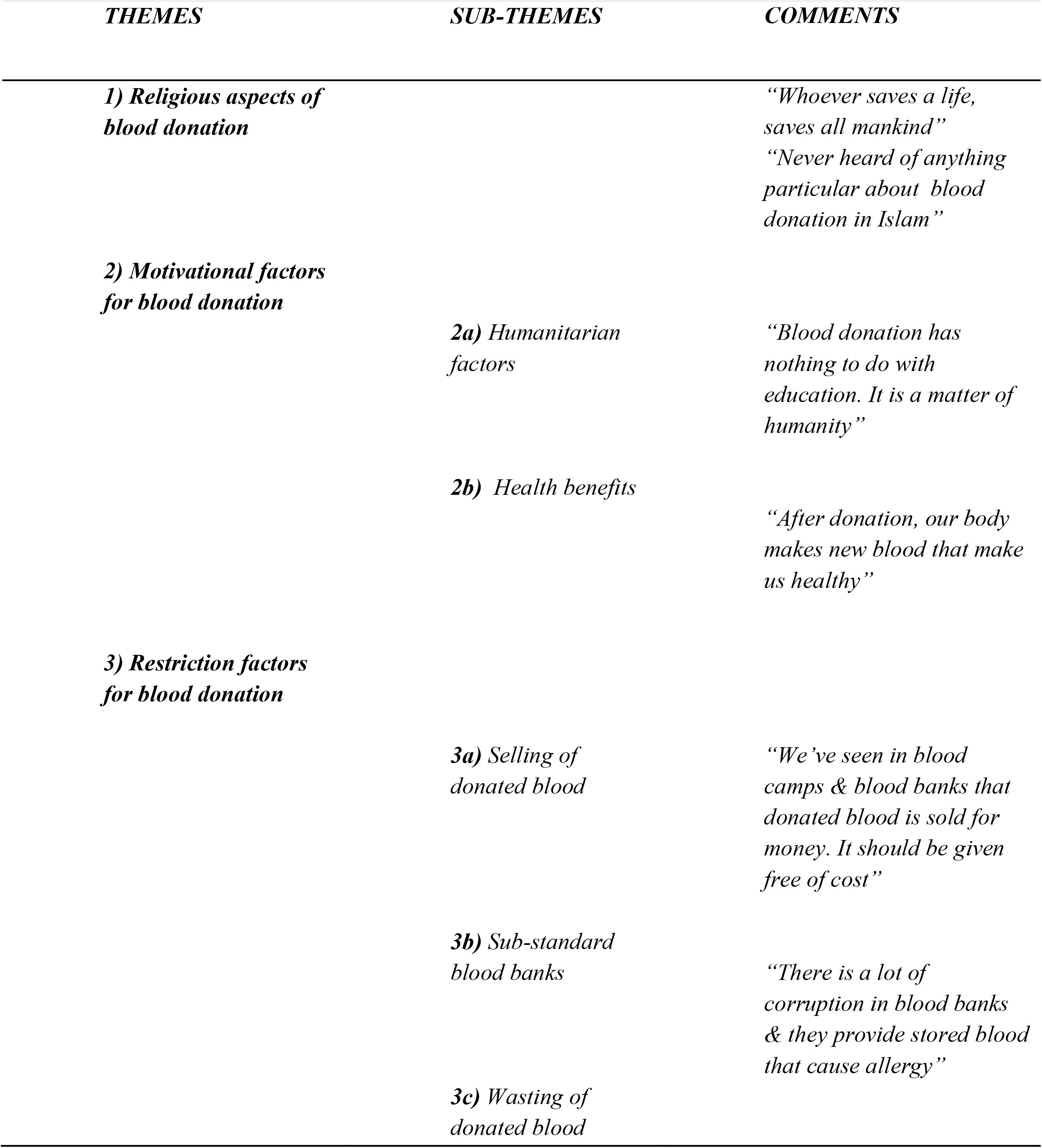
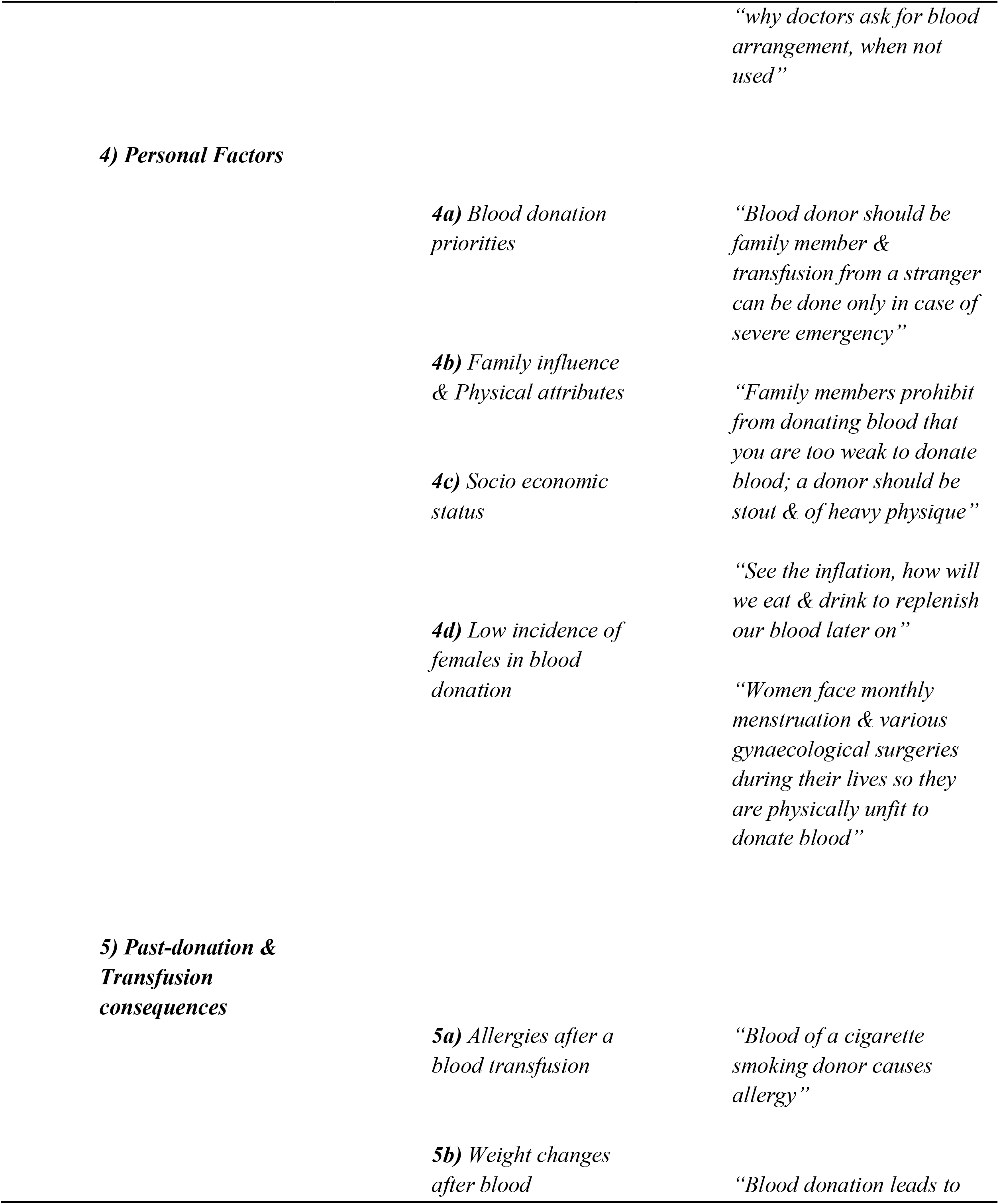
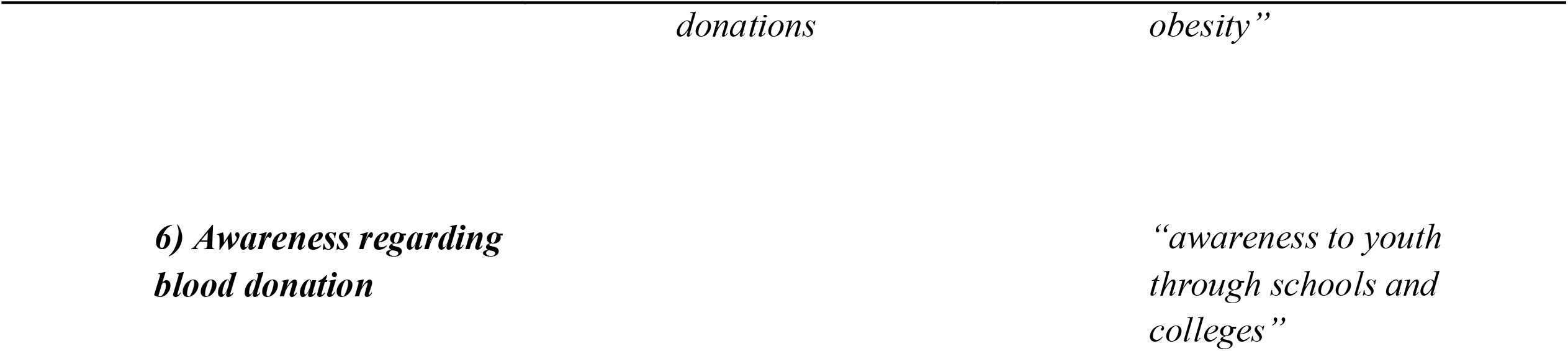
Themes and Subthemes The summary of results is given in Table 1.

## DISCUSSION

In Pakistan, the demand for blood donation is extremely high, and it is difficult to maintain the flow of safe supplies. According to 2022 Revision of World Population Prospects, median age in Pakistan is 20.6 years which is theoretically adequate to fulfil blood demand [10]. However, a lack of knowledge, cultural barriers, misunderstandings, and inadequate infrastructure hinder blood supply chains.

The restricted supply of safe blood poses threat healthcare quality. Therefore, the present study aimed to rule out not only major perceptions towards blood transfusion but also their link to religion, cultural background, personal behaviors, social influence, and poverty.

Religion is firmly ingrained in Pakistani society, and is a primary motivator for blood donation. In the current survey, participants were of the view that helping others is a religious obligation. It was discovered concurrently with a study in Bangladesh [11]. However, in a study in Nigeria, people were hesitant to receive blood due to their religious beliefs [12]. In a nutshell, Religion has a substantial impact on blood donation rates, both positively and negatively. If we want to design a targeted public awareness campaign, the basic role of religion cannot be emphasized.

Also, some personal experiences played a significant part in determining the intention to donate blood. Predominant among all were beneficial health effects on the body due to blood regeneration. Probably due to modification in lipid profile after donation as in a study by Al haji Bukar [13]. Other advantages after blood donation include contentment, improved attentiveness, and better well-being [14, 15].

Moving on, another key factor is society, particularly family and friends. They influence donors and non-donors to donate as in a study by McCombie on Blood donation patterns of undergraduate students [16]. Because the transmission of values from generation to generation among family members who donate blood, as well as the influence of active blood donors on others, is undeniable [17]. However, in our study, family played a role in both driving and discouraging voluntary donation.

Even if religious, personal, and social forces are all aligned to drive a person to donate, a donor can still encounter pushback in the form of poor blood bank management in Pakistan. Selling and wastage of donated blood, misguidance about donation, and unsafe screening practices [18] halt the supply chain. The majority of the blood banks in Karachi as well as all across Pakistan had practices that were much below WHO guidelines [19]. Participants proposed that advanced screening needs to be done in the Blood banks of Pakistan. It is a major restricting factor.

Nearly 9 million people in Pakistan live below the poverty line. Low socio-economic background restricts people from donating even if they are eligible. Because they require more nourishment to compensate for the blood loss caused by donation. Due to poverty, they are unable to buy healthy food. Hence, the number of voluntary unpaid transfusions can increase as people’s access to improved life opportunities increases. According to the WHO, developed countries produce more blood donors than developing and low-income countries because improved living conditions make it easier for people to donate blood [4].

The blood donor pools did not adequately reflect the female population, whereas women makes up more than half of the entire population of Pakistan. The majority thought that women were less able to donate blood because they experienced more blood insufficiency than men. Most participants in this survey claimed that they had never been asked for a donation. However, a major barrier to blood donation is the lack of knowledge that women can donate if their Hb is maintained through the use of iron supplements or another method. For instance, 92.7% of the donors in the New York blood center who were rejected owing to low hemoglobin levels were female [20].As a result, and in line with several studies, we can increase the number of female donors by giving out iron supplements together with suitable and tailored monitoring of Blood Hb-level [21].

One noteworthy feature of this study was how participants believed that blood donation contributed to obesity. A Malaysian study revealed that the link between blood donation and fat is a myth. One possible reason for this could be that individuals eat frequently after donating blood, which could lead to weight gain. Otherwise, solely donating blood does not cause obesity [22].

Blood Transfusion and Donation are both very risky procedures even after screening [23]. In the current study, participants expressed concern over post-transfusion reactions. Such reactions reduced the pool of potential blood donors, and Patients may choose direct donation from relatives or friends over “strangers” because they believe it minimizes the possibility of transfusion transmissible infections [24].

Lastly, participants in the current study were not adequately informed on the benefits and hazards of blood donation and blood transfusion in general, thus the broad idea of blood donation remains inadequate. Another study conducted in Pakistan found that half of the recipients was unwilling to pay extra for blood screening and that they were unaware of its significance. This lack of awareness has contributed to poor health effects. [25]

All the parameters emphasize the importance of awareness about blood donation. One possible way of raising awareness is to consider the behavior change pathway guidance given by the World Health Organization (WHO). In this report, they centered on the importance of increased awareness and education among the general public. It was also underlined that a combination of policy tools, such as law, regulation, tobacco control, etc., might enhance donations. [26]

In Additional educational plans are required to raise awareness. The majority of people thought social media was helpful. In an effort to increase blood donation awareness, Facebook began working on a blood donation feature in 2018. A US study found that blood donations rose by 4.0% and donations from new donors by 18.9% when people used Facebook’s blood donation tool. [27]

Some participants suggested including in school curricula the essential knowledge of blood transfusion. Consequently, active health programmes on blood donation and transfusion should be introduced through various media, including the internet. Cristiano Ronaldo, the well-known international football player, and the National Health Ministry’s Safe Blood Transfusion Programme inked a deal to advance the voluntary blood donation culture in Pakistan. [28]

These can urge healthy Pakistanis, as well as their family members and friends, to donate blood as frequently as possible in order to ensure that blood is accessible for all patients in need. This study has clarified key aspects like religion, personal beliefs, social circle, socioeconomic level, and challenges due to blood bank mismanagement that play a part in determining an individual’s tendency to donate and consequently, the size of blood pool nationally.

### Limitations of the Study

This information is helpful for establishing blood donation strategies in the future and for inspiring young people. Despite this, it has certain limitations particularly with regard to sampling because it only covered participants from 2 units of a hospital.Individuals from various institutes, both public and private, should be investigated for the same subject to validate the results and extend the overall picture.

## Conclusion

This study uncovered the flaws in Pakistan’s blood donation system as well as the perceptions of those who donate or could donate blood. The need of the hour is to develop initiatives to urge current donors to donate more frequently and to encourage others who are eligible to start donating. Although responses on the motivational and limiting components of blood donation indicate the importance of family and friends, religion, personal values, and economic status, more research is required. Misinformation at blood banks, blood selling, fear of allergic reactions, and a low socioeconomic background are all issues that prevent people from donating blood.

## Data Availability

All data produced in the present work are contained in the manuscript

